# Familial clustering of erosive hand osteoarthritis in a large statewide cohort

**DOI:** 10.1101/2020.04.04.20053264

**Authors:** Nikolas H. Kazmers, Huong D. Meeks, Kendra A. Novak, Zhe Yu, Gail L. Fulde, Joy L. Thomas, Tyler Barker, Michael J. Jurynec

## Abstract

**Objectives:** Erosive hand osteoarthritis (EOA) is a severe and rapidly progressing form of osteoarthritis. Its etiology remains largely unknown, which has hindered development of successful treatments. Our primary goal was to test the hypothesis that EOA would demonstrate familial clustering in a large statewide population linked to genealogical records, which would suggest a genetic contribution to the pathogenesis of this condition. Our secondary purpose was to determine the association of potential risk factors with EOA.

**Methods:** Patients diagnosed with EOA were identified by searching medical records from a comprehensive statewide database, the Utah Population Database (UPDB). Affected individuals were then mapped to pedigrees to identify high-risk families with excess clustering of EOA as defined by a Familial Standardized Incidence Ratio (FSIR) of ≥ 2.0. The magnitude of familial risk of EOA in related individuals was calculated using Cox regression models. Association of potential EOA risk factors was analyzed using conditional logistic regression and logistic regression models.

**Results:** We identified 703 affected individuals linked to 240 unrelated high-risk pedigrees with excess clustering of EOA (FSIR ≥ 2.0). The relative risk of developing EOA was significantly elevated in first-degree relatives. There was a significant association with the diagnosis of EOA and age, sex, diabetes, and obesity.

**Conclusions:** Familial clustering of EOA observed in a statewide database indicates a potential genetic contribution to the etiology of the disease. Identification of causal gene variants in these high-risk families may provide insight into the genes and pathways that contribute to EOA onset and progression.

## Introduction

Hand osteoarthritis (OA) is the most prevalent form of OA and is a major cause of disability.^1-7^ It is a heterogeneous disorder with a substantial genetic contribution. ^8^ Despite significant heritability of hand OA, very few genes and pathways have been discovered that modify the course of the disease.^6, 7^ Erosive hand osteoarthritis (EOA) is considered a more severe form of hand OA that affects the distal and proximal interphalangeal joints.^9-13^ EOA is defined by its sudden onset, rapid progression, and radiological evidence of central subchondral erosions that have a ‘gull-wing’ or ‘saw-tooth’ appearance, collapse of the subchondral bone, and marginal osteophyte formation.^9, 14, 15^ Despite the prevalence and severity of EOA^16, 17^, there are no therapeutics that are effective in preventing the onset or limiting the progression of the disease.^18^The main obstacle to the development of disease-modifying therapies is limited understanding of the disease process.^19, 20^ We have limited knowledge of the genes that confer susceptibility to EOA.^21-23^ EOA is suggested to have a familial contribution^24^, but this analysis was limited to sibling pairs. The genetic studies of EOA to date have been limited in size and scope, which has hindered to identification of targets for development of therapeutic intervention.

There have been several described risk factors associated with EOA including sex, alcohol consumption, and obesity, although there has been some discrepancy in risk factors between cohorts.^16, 17, 25, 26^ Determining the contribution of risk factors in different cohorts allows for a more representative view of patient characteristics associated with the pathogenesis of EOA, and may provide clinically useful information to identify groups at an increased risk of disease development.

Our goal was to utilize the Utah Population Database (UPDB), a large statewide population database linked to comprehensive genealogical records,^27-34^ to perform a retrospective population-based study to i) test our hypothesis that EOA clusters in large families, ii) define the magnitude of familial risk of EOA, and iii) evaluate our cohort for potential risk factors associated with EOA.

## Methods

### Study Approval

This study was approved by the Institutional Review Boards of the University of Utah (IRB # 79442) and Intermountain Healthcare (IRB # 1050554) and by the Resource for Genetic and Epidemiologic Research.

### The Utah Population Database (UPDB)

Our study utilizes data drawn from UPDB (https://uofuhealth.utah.edu/huntsman/utah-population-database/). The UPDB is one of the world’s largest and most comprehensive sources of linked population-based information for demographic and genetic studies. The UPDB contains data on over 11 million individuals from the late 18th century to the present. UPDB data represent Utah’s population that appear in administrative records. Data are updated as they become available from statewide birth and death certificates, hospitalizations, ambulatory surgeries, and drivers licenses. UPDB creates and maintains links between the database and the medical records held by the two largest healthcare providers in Utah as well as Medicare claims. The multigenerational pedigrees representing Utah’s founders and their descendants were constructed based on data provided by the Genealogical Society of Utah (GSU). Pedigrees spanning the past century have been expanded extensively based on vital records and, together with the GSU data, form the basis of the deep genealogical structure of the UPDB. The UPDB has been used in the early investigational stages to demonstrate familial clustering of diseases^35-37^, and has been instrumental to the discovery of many disease causing genes, including breast and ovarian cancer^31, 34^, colon cancer^28^, and prostate cancer.^38^

### Selection of Cases

We identified individuals diagnosed with erosive hand osteoarthritis (EOA) between October 1^st^, 2015 - December 31^st^, 2019 in the UPDB using the International Classification of Diseases (ICD) Tenth revisions code: ICD-10 M15.4. Individuals were excluded if they were also diagnosed with rheumatoid arthritis (ICD-9 714.0, ICD-10 M05.xx), other rheumatoid arthritis subtypes (ICD-9 714.2, ICD-10 M06.xx), or juvenile rheumatoid arthritis (ICD-9 714.3, ICD-10 M08.xx). Affected individuals were required to have relatives in the UPDB to be included in our study cohort so we could link them to pedigrees.

### High Risk Pedigree Identification

To determine if there was excess familial clustering of EOA in each pedigree, we utilized the Familial Standardized Incidence Ratio (FSIR).^39^ FSIR allows for the quantification of familial risk of a disease by comparing the incidence of a disease in a family to its expected incidence in the general population. FSIR is a statistical method that accounts for the number of biological relatives in a pedigree, the degree of relatedness, and the age at which an individual is diagnosed. Exact one-sided Poisson probabilities were calculated under the null hypothesis of no familial enrichment of EOA. Individuals were grouped into fourteen categories based on age (0-30, 31-40, 41-50, 51-60, 61-70, 71-80, and 81-120) and sex. To determine the incidence ratio, the number of years prior to and after diagnosis was calculated for all affected and unaffected individuals, and then the number of living diagnosed years was divided by the number of living undiagnosed years. To determine the pedigree incidence ratio, the UPDB was analyzed to identify the founders of pedigrees containing an affected individual, the affection status of every individual biological relative in each pedigree was determined, and incidence ratio was calculated as described above. The pedigree’s incidence ratio/whole population incidence ratio was used to determine the FSIR. High-risk pedigrees were selected if they had two or more living affected individuals, and if the FSIR was ≥ 2 and significant (p < 0.05) using a chi-squared test as described by Kerber.^39^

### Familial Risk Analysis

Controls were matched to cases in a 10:1 ratio by sex, birth year, and whether born in Utah. Additionally, we imposed the restrictions that controls must be alive in the matched cases’ diagnosis year. Cases and controls were followed from birth until death, or 2019, or diagnosis year, whichever occurred first. Estimates of familial risk were based on a hazard rate ratio (HR) of familial recurrence, which represents the ratio of the hazard rate for the occurrence of EOA among relatives of the cases with the comparable hazard rate among the relatives of the matching controls.

The HRs were calculated using Cox regression models, additionally adjusting for sex and birth year. Because observations within families are non-independent, a Huber-White sandwich estimator of variance for clustered data was used to correct for any families that were analyzed multiple times because of the multiple EOA cases within the family.^40^ Analyses were performed separately in which specific groups of relatives of the cases were compared to the comparable relatives of the matched controls as follows: first degree relatives, second degree relatives, first cousins and second cousins.

### Age-standardized sex-specific EOA incidence rates

We selected all individuals with birth year and informative sex who resided in Utah from 2015 until 2018 or died in Utah, whichever happened first. This resulted in identification of 603 individuals with EOA. In contrast to the EOA cohort used to determine familial risk and identification of high-risk pedigrees, we chose to exclude the patients diagnosed with EOA in 2019 because 2018 is the last year the UPDB received death certificates. Demographic characteristics of the EOA cases and non-EOA population were compared using t-tests for continuous variables and chi-square tests for categorical variables (Supplemental Table 1).

**Table 1.** Baseline Patient Characteristics of the Erosive Hand Osteoarthritis Cohort.

|  |  |
| --- | --- |
| Number of Individuals | 703 |
| Age (years) | 67 ± 11.54 (Range 12-94) |
| Race |  |
| White | 639 (90.9%) |
| Non-white | 64 (9.1%) |
| Sex |  |
| Female | 564 (80.23%) |
| Male | 139 (19.77%) |

Age-standardized incidence rates by sex were calculated using the direct method. Person-years were calculated for affected individuals (cases) and unaffected individuals (controls). Cases contributed one person-year for every year lived in Utah from 2015 until diagnosed with EOA. Person-years contributed from each control was one additional year for every year lived in Utah from 2015 until death or 2018, whichever occurred first. The female-to-male incidence ratios were calculated by dividing the rate in males by that in females for each age group, and the corresponding 95% CI was estimated assuming log-normal distribution. Logistic regression models were used to assess the association between EOA and sex, additionally adjusting for birth year and whether the subjects were Caucasian and Hispanic.

### Risk Factor Analysis

Specific ICD-9 and ICD-10 codes were used to identify risk factors among study patients (Supplemental Table 2). Relative risk of EOA were calculated using gender-specific Cox proportional hazards models with adjustments for sex, birth year, race and ethnicity, and clustering for common mothers. We adjusted independently for obesity and diabetes since obesity is a risk factor for type 2 diabetes. Follow up time as defined by year from 2015 to date of death, date of last reside in Utah, or date of first EOA diagnosis, whichever happened first. Odds ratios and 95% CI were calculated.

**Table 2.** High-Risk Pedigrees with Excess Familial Clustering of Erosive Hand Osteoarthritis.

| Founder Birth Year | Number of Descendants | Number of Affected Individuals | FSIR<sup>#</sup> |
| --- | --- | --- | --- |
| 1682* | 109,720 | 15 | 2.1 |
| 1789 | 81,204 | 12 | 2.1 |
| 1758 | 34,750 | 8 | 3.5 |
| 1795 | 300,10 | 8 | 3.0 |
| 1715 | 38,586 | 8 | 2.4 |
| 1780 | 34,156 | 7 | 2.8 |
| 1794 | 7,262 | 5 | 9.7 |
| 1779 | 13,015 | 5 | 4.7 |
| 1762 | 6677 | 4 | 11.8 |
| 1805 | 5,182 | 3 | 15.5 |

Abbreviations: FSIR = familial standardized incidence ratio.
\* indicates pedigree represented in Figure 1.

## Results

### Identification and demographic detail of the erosive hand osteoarthritis cohort

To identify individuals diagnosed with EOA, we searched the UPDB for individuals with the ICD-10 code ICD-10 M15.4 from October 2015 – December 2019 and excluded patients with a rheumatoid arthritis diagnosis. This query identified 703 individuals for analysis with a mean age at time of diagnosis was 67 years (± 11.54), 80.23% were female, and 90.9% of individuals were white (Table 1).

### Identification of High-Risk Pedigrees

To test if there is significant familial clustering of EOA in our cohort, we analyzed individuals diagnosed with EOA that linked to a pedigree using the Familial Standardized Incidence Ratio (FSIR) calculation.^39^ We identified 240 unrelated, multigenerational, high-risk pedigrees that had at least two living members in the UPDB and an increased clustering of EOA, defined by a FSIR ≥ 2.0 (p-value < 0.05). Of the 240 high-risk pedigrees, the FSIR ranged from 2.0 – 210.8 (mean 11.7 ± SD = 11.7 ± 210.2; 1st and 3rd quartiles = 4.8 and 11.7, respectively). Founder birth year, number of descendants, number of affected individuals, and FSIR values are indicated for 10 representative high-risk pedigrees in Table 2. Figure 1 is an example of a multigenerational high-risk pedigree with at least 15 known affected individuals and a FSIR of 2.06. The identification of high-risk pedigrees indicates significant familial clustering of EOA in our cohort.

**Figure 1.**
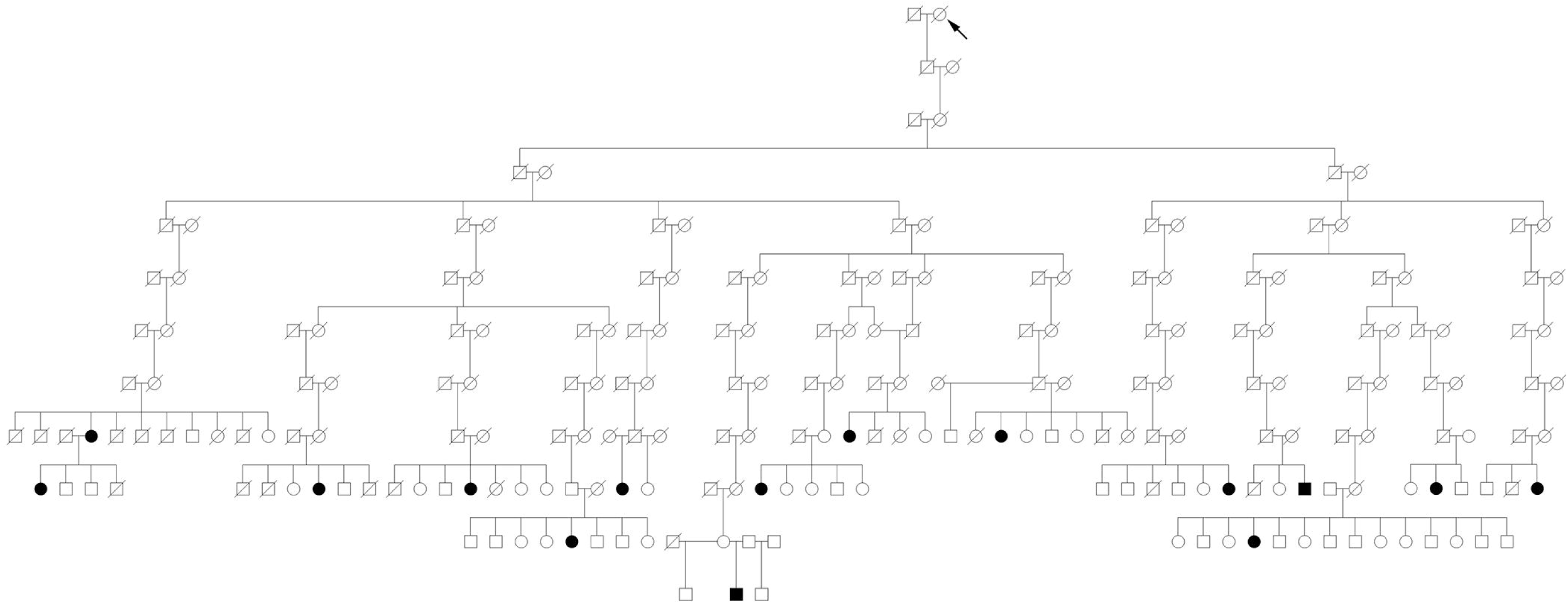
Example of a high-risk erosive hand osteoarthritis pedigree identified in the Utah Population Database. Circles = females, squares = males, arrow = family founder, slashes = deceased. White circles/squares = affection status unknown. Black circles/squares = individuals affected with erosive hand osteoarthritis.

### Familial Risk

To determine whether there is an increased risk of EOA among closely related individuals, we examined the relative risk of developing EOA in first and second-degree relatives and first and second cousins in our cohort. The risk of developing EOA was approximately 5.5-fold greater in first-degree relatives of EOA cases compared to controls (Relative Risk, 5.53 [95% CI, 2.1 - 14.58], p < 0.001) (Table 3). We were unable to detect a significant elevated risk of EOA in second-degree relatives or first and second cousins of EOA cases. Together with the familial clustering of EOA, these data suggests an underlying genetic contribution to EOA.

**Table 3.**
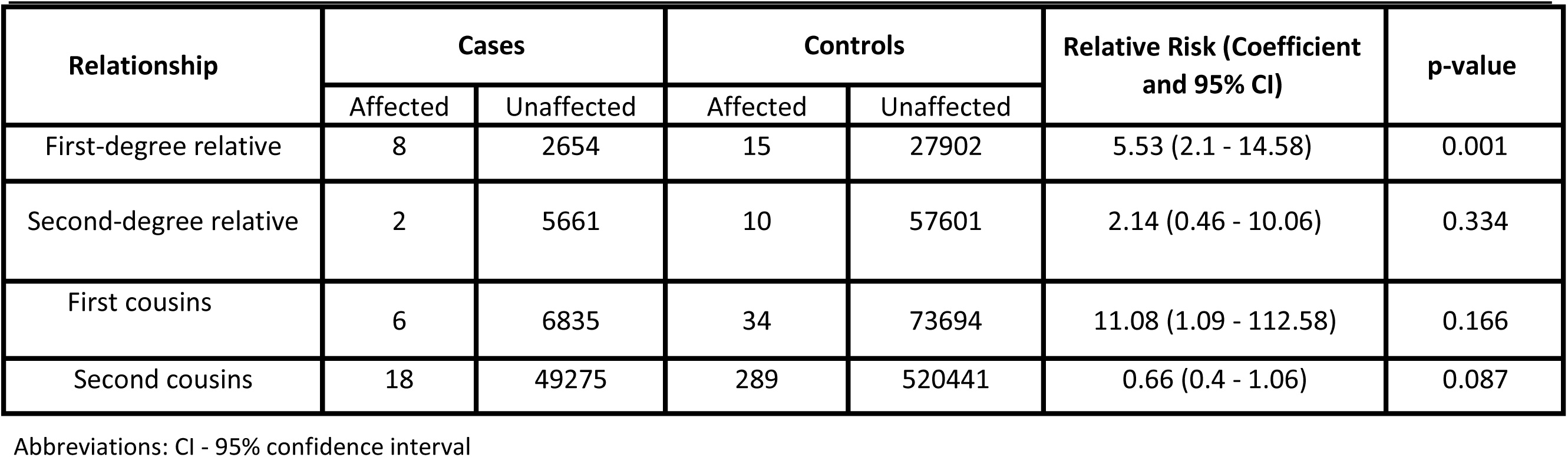
Increased Familial Risk of Erosive Hand Osteoarthritis.

| Relationship | Cases |  | Controls |  | Relative Risk (Coefficient and 95% CI) | p-value |
| --- | --- | --- | --- | --- | --- | --- |
|  | Affected | Unaffected | Affected | Unaffected |  |  |
| First-degree relative | 8 | 2654 | 15 | 27902 | 5.53 (2.1 - 14.58) | 0.001 |
| Second-degree relative | 2 | 5661 | 10 | 57601 | 2.14 (0.46 - 10.06) | 0.334 |
| First cousins | 6 | 6835 | 34 | 73694 | 11.08 (1.09 - 112.58) | 0.166 |
| Second cousins | 18 | 49275 | 289 | 520441 | 0.66 (0.4 - 1.06) | 0.087 |

Abbreviations: CI - 95% confidence interval

### Age-Standardized Sex-Specific Incidence Rates of EOA

Hand OA affects females more than males and this trend appears to be valid for EOA.^6, 7, 13, 24, 25^ To determine if there is an age and sex bias associated with EOA, we examined age-standardized sex-specific incidence rates of EOA in our statewide cohort from October 2015 - December 2018. We found a significant association between sex and age with EOA. Out of 2,065,277 controls and 606 EOA cases, 80% of EOA cases were female (51.5% female in the controls) and older (birth year, EOA cases-1950.7 ± 11.5 and controls - 1979.8 ± 23) when compared to controls (p < 0.001) (Supplemental Table 1). We also determined that females have a significantly higher rate of EOA from the ages of 40-89 compared to males, with the highest female-to-male incidence ratios being 4.730 (95% CI, 3.956 – 5.655) in the 60-69 age group (Table 4). Logistic regression analysis indicated that females have a 3.48-fold increased risk of EOA diagnosis compared to males even after adjusting for birth year, race and ethnicity (Relative Risk, 3.48 [95% CI, 2.85 – 4.25). Our results indicate that being female is a significant risk factor for EOA.

**Table 4.**
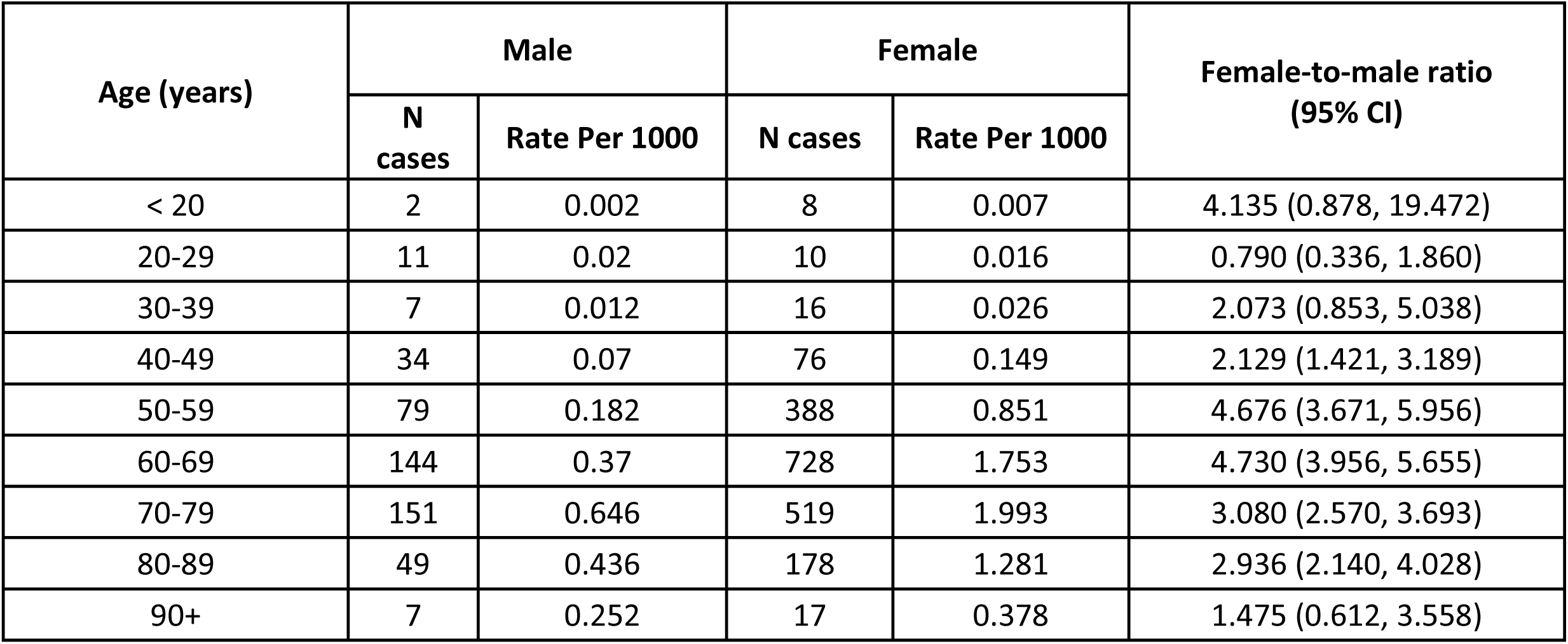
Age-Specific Incidence Rate of EOA by Sex and Female-to-Male Incidence Ratios.

| Age (years) | Male |  | Female |  | Female-to-male ratio<br>(95% CI) |
| --- | --- | --- | --- | --- | --- |
|  | N cases | Rate Per 1000 | N cases | Rate Per 1000 |  |
| < 20 | 2 | 0.002 | 8 | 0.007 | 4.135 (0.878, 19.472) |
| 20-29 | 11 | 0.02 | 10 | 0.016 | 0.790 (0.336, 1.860) |
| 30-39 | 7 | 0.012 | 16 | 0.026 | 2.073 (0.853, 5.038) |
| 40-49 | 34 | 0.07 | 76 | 0.149 | 2.129 (1.421, 3.189) |
| 50-59 | 79 | 0.182 | 388 | 0.851 | 4.676 (3.671, 5.956) |
| 60-69 | 144 | 0.37 | 728 | 1.753 | 4.730 (3.956, 5.655) |
| 70-79 | 151 | 0.646 | 519 | 1.993 | 3.080 (2.570, 3.693) |
| 80-89 | 49 | 0.436 | 178 | 1.281 | 2.936 (2.140, 4.028) |
| 90+ | 7 | 0.252 | 17 | 0.378 | 1.475 (0.612, 3.558) |

### Risk Factors Associated with EOA

Knowledge of risk factors that may contribute to EOA remains incomplete. We analyzed the association of several risk factors with EOA that have been previously associated with general hand OA and EOA (see Supplemental Table 2 for risk factor diagnostic codes).^16, 17, 25, 26^ We examined the association of tobacco use, alcohol use, diabetes, obesity, and having a first-degree relative with EOA in the same cohort used for the age- and sex-specific analysis. Because females were at a higher risk for EOA (Table 4), we examined the relative risk of the above risk factors independently in males and females while adjusting for demographic features (see Methods and Supplemental Table 1). A history of obesity (Relative Risk, 1.50 [95% CI, 1.18 – 1.90]) was significantly associated with EOA in females, while diabetes was significantly associated in males (Relative Risk, 1.65 [95% CI, 1.05 – 2.59]). In both the female and male EOA cohorts, independent of obesity and diabetes, having a first-degree was a significant risk factor for EOA with the risk being greater in males than females (Table 5). No significant associations were detected between EOA diagnosis and tobacco or alcohol use. These data indicate obesity, diabetes, and having a first-degree relative with EOA are all significant risk factors for EOA.

**Table 5.** Risk Factors Associated with Erosive Hand Osteoarthritis.

| Risk Factor/Clinical Diagnosis<br>(not adjusted for obesity) | Relative Risk with<br>95% Confidence<br>Interval | p-value |
| --- | --- | --- |
| **Females** |  |  |
| Tobacco Use | 1.3 (1.00 – 1.68) | 0.051 |
| Alcohol Use | 0.80 (0.26 – 2.51) | 0.707 |
| Diabetes | 1.23 (0.95 – 1.60) | 0.114 |
| FDR* with EOA | 6.40 (2.65 – 15.43) | < 0.001 |
| **Males** |  |  |
| Tobacco Use | 0.79 (0.48 – 1.30) | 0.361 |
| Alcohol Use | 2.48 (0.93 – 6.62) | 0.069 |
| Diabetes | 1.65 (1.05 – 2.59) | 0.031 |
| FDR with EOA | 16.99 (5.37 – 53.75) | < 0.001 |

| Risk Factor/Clinical Diagnosis<br>(not adjusted for diabetes) | Relative Risk with<br>95% Confidence<br>Interval | p-value |
| --- | --- | --- |
| **Females** |  |  |
| Tobacco Use | 1.27 (0.98 – 1.65) | 0.075 |
| Alcohol Use | 0.80 (0.26 – 2.50) | 0.704 |
| Obesity | 1.50 (1.18 – 1.90) | 0.001 |
| FDR with EOA | 6.42 (2.66 – 15.49) | < 0.001 |
| **Males** |  |  |
| Tobacco Use | 0.82 (0.49 – 1.35) | 0.426 |
| Alcohol Use | 2.48 (0.93 – 6.64) | 0.069 |
| Obesity | 1.22 (0.66 – 2.24) | 0.526 |
| FDR with EOA | 16.92 (5.35 – 53.54) | < 0.001 |

\* FDR = first-degree relative.

## Discussion

We have used a unique statewide medical genetics resource, the Utah Population Database (UPDB), to identify a cohort of individuals diagnosed with erosive hand osteoarthritis (EOA). From this cohort we have i) identified 240 unrelated high-risk pedigrees demonstrating familial enrichment EOA, ii) determined that first-degree relatives of an individual with EOA is at approximately 5.5-fold increased risk of developing EOA, and iii) that sex, age, diabetes, obesity, and having a first-degree relative with EOA are significant risk factors associated with EOA. In sum, these data suggest that both genetic and physiological factors contribute to the development of EOA in a large population-based cohort.

### Genetic Involvement in EOA

Although hand OA is highly heritable^8^, few genes with large effects have been associated with the onset and progression of hand OA^6, 7^, and only three genes have been associated with the EOA phenotype.^21-23^ The predominant approach to discover hand OA gene variants has been genome-wide association studies (GWAS)^41-46^, which relies on large cohorts of cases and controls and well-defined phenotypes. The heterogeneous nature of hand OA has likely been a confounding factor in some GWAS. An alternative approach to GWAS is to study families with highly penetrant, severe or early-onset forms of OA.

The study of rare variants in affected families is a powerful way to identify gene variants with a determinate effect on disease development.^47-50^ Using the UPDB, we have identified 240 large multigenerational, high-risk pedigrees segregating EOA as an apparent dominate trait. Although a previous study described association of EOA in sibling pairs^24^, our study is the first to identify a large number of multigenerational EOA pedigrees and determine relative risk among family members. Identification of causal gene variants in these families will inform us about genes and pathways that when disrupted contribute to EOA.

### Risk Factors Associated with EOA

We examined the risk of developing EOA based on sex and age and found that females are 3.48-fold more likely to develop EOA than males with the highest female-to-male incidence ratios in the 60-69 age group. This suggests that EOA is similar to general hand OA in that females are disproportionately affected.^6, 7, 13, 24, 25^ When we subdivide risk factors based on sex and adjusted for a family history of EOA and other demographic factors, we found that obesity was a risk factor in females and diabetes a risk factors in males, while having a first-degree relative with EOA is a common risk factor to both sexes. Our data are consistent with other studies that have examined risk factors for EOA in other populations.^16, 17, 25, 26^ Awareness of these comorbidities observed to be significantly associated with EOA in the current study may help guide the clinical diagnosis of this condition in at-risk populations.

This study has several limitations. As for all database studies, it is unclear how errors in diagnostic coding would impact the study findings, and manual chart review was not possible for all individuals included in the analysis. Prior investigation has shown 93-97% rates of accuracy for UPDB diagnostic coding when compared to manual chart review.^51, 52^ The relative risk and FSIR calculations are likely underestimates for EOA, which is due to several factors. Our cohort was limited to individuals with an ICD-10 diagnosis for EOA, which has only been in use since October 2015, and our high-risk pedigree analysis can only identify individuals that have been diagnosed in Utah. We are missing individuals who were diagnosed using different codes prior to October 2015 and those that were diagnosed out of state. Because of this, in high-risk pedigrees we consider individuals without an EOA diagnosis as ‘affection status unknown’ until we can definitively determine if they are unaffected or affected. Our study does not evaluate the extent to which EOA is genetic. The enrichment of EOA in pedigrees is suggestive of a genetic contribution, especially in distant relatives, but we cannot rule out outer factors such as environmental influence on EOA particularly in light of the risk factor associations we observed.

To conclude, we demonstrated that EOA demonstrates familial enrichment, an increased relative risk among first-degree relatives, and identified significant EOA risk factors. Taken together, these findings suggest that EOA has a genetic and environmental component to its etiology. Genomic analysis of affected and unaffected individuals within our high-risk pedigrees holds promise in identifying genetic variants associated with EOA. By identifying and studying gene variants that cause EOA, we may learn about the biological mechanisms that lead to other forms of OA, which may provide significant insight into surgical treatment or therapeutic intervention.

## Data Availability

All data is available upon reasonable request.

## Acknowledgments and affiliations

This study was funded by the Skaggs Foundation for Research and the Utah Genome Project (M.J.J. and N.H.K.). We thank Dr. Rena D’Souza for critical feedback. We acknowledge the Pedigree and Population Resource of the Huntsman Cancer Institute for its support of the UPDB. We also acknowledge partial support for the UPDB through grant P30 CA2014 from the NCI and from the University of Utah’s Program in Personalized Health and Center for Clinical and Translational Science.

---

**Supplemental Table 1** – Study Population Used for Age-Standardized Sex-Specific Incidence Rates of Erosive Hand Osteoarthritis.

**Supplemental Tale 2** – Identification of Risk Factors Using Diagnostic Coding.

## Key messages

### What is already known about this subject?

1. Erosive hand osteoarthritis (EOA) is a serve, rapidly progressing form of osteoarthritis. Previous studies have identified several associated risk factors in EOA cohorts.

### What does this study add?

1. Our study indicates that EOA can cluster in large, multigenerational families. First-degree relatives of an individual diagnosed with EOA has an elevated risk of developing the EOA, suggesting a genetic contribution to the disease.
2. Risk factor analysis indicates that age, sex, and obesity are risk factors for females, whereas diabetes is a risk factor for males. Having a first-degree relative was a major risk factor in both sexes. Tobacco and alcohol use are not risk factors in our cohort. How might this impact on clinical practice or future developments?
3. Our findings may guide the clinical diagnosis and treatment of EOA in at-risk populations.
4. Genetic analysis of our high-risk EOA pedigrees will allow for the identification of casual gene variants, which may inform rational therapeutic development to prevent or slow the progression of EOA.

## Notes

### Competing Interest Statement

The authors have declared no competing interest.

